# Spatio-temporal distribution of rhinovirus types in Kenya: A retrospective analysis, 2014

**DOI:** 10.1101/2024.04.02.24305207

**Authors:** John Mwita Morobe, Everlyn Kamau, Martha M. Luka, Nickson Murunga, Clement Lewa, Martin Mutunga, Godfrey Bigogo, Nancy Otieno, Bryan Nyawanda, Clayton Onyango, D. James Nokes, Charles N. Agoti, Patrick K. Munywoki

## Abstract

Rhinoviruses (RVs) are one of the most commonly detected viruses in people with acute respiratory illness (ARI). Despite their significant disease burden, RV epidemiology at national levels is underexplored. The circulation patterns of RV types throughout a population and the role of virus genotype in this distribution are ill-defined. We generated 803 VP4/VP2 gene sequences from rhinovirus-positive samples collected from ARI patients, including both in-patient and outpatient cases, between 1^st^ January and 31^st^ December 2014 from eleven surveillance sites across Kenya and used phylogenetics to characterise virus introductions and spread. RVs were detected throughout the year, with the highest detection rates observed from January to March and June to July. We detected a total of 114 of the 169 currently classified types. Our analysis revealed numerous virus introductions into Kenya characterized by local expansion and extinction, and extensive spatial mixing of types within the country due to the widespread transmission of the virus after an introduction. This work demonstrates that in a single year, the circulation of rhinovirus in Kenya was characterized by substantial genetic diversity, multiple introductions, and extensive geographical spread.

## Introduction

Rhinoviruses (RVs) are one of the leading cause of acute upper respiratory tract illnesses, commonly referred to as “common cold”^1, 2^. RV infections occur all year, although they peak in early autumn and late spring in many temperate countries, and in the rainy seasons in tropical countries^2, 3^. RVs cause 8 – 12 episodes of respiratory infections in children and 2 – 4 episodes in adults each year ^4–6^, posing significant social and economic burden due to time lost from work or school, medical attendance, and reduced performance of regular duties^7^. Despite the high prevalence and clinical importance of RV infections, little is known about the patterns of occurrence of RV types, as well as the role of virus type in the RV distribution.

RVs belong to the genus *Enterovirus* of the family *Picornaviridae*. The virus genome consists of ∼7200 nucleotides and encodes four structural proteins (VP4, VP2, VP3 and VP1) and seven non-structural proteins (2A, 2B, 2C, 3A, 3B, 3C and 3D)^2^. The single-stranded positive sense RNA genome is enclosed within a protein capsid that is made up of sixty copies of each of the four structural proteins, of which VP1, VP2, and VP3 are exposed outside, while VP4 is completely masked in the capsid^2, 8, 9^. The three surface-exposed capsid proteins carry the antigenically important sites^2, 8, 9^. Genetic variation in the VP4/VP2 and VP1 genomic regions have been useful in molecular typing^10, 11^ and epidemiological investigations^12–14^. As of 15^th^ March 2023, a total of 169 RV types have been described and classified into three distinct species, i.e., *Rhinovirus A, Rhinovirus B,* and *Rhinovirus C*^15^.

Viral sequence data are increasingly used to track the geographic spread and transmission of viral pathogens^16, 17^. Genomic data has considerably informed public health interventions and outbreak management for viral pathogens, e.g., SARS-CoV-2, Ebolavirus, and Zika virus^18–21^. Widespread spatial and temporal transmission patterns of RV was previously described within the Kilifi Health and Demographic Surveillance System (KHDSS) on the Kenyan Coast covering around 300,000 residents^22^ and marked by multiple virus introductions^12^. Transmission patterns of RV at a nationwide scale have not been documented in Kenya. Here we analyze 803 sequences obtained from individuals presenting with acute respiratory illness (ARIs) at 11 sentinel surveillance sites across Kenya in 2014 ^23^, to explore the temporal and spatial circulation patterns of RV types at the countrywide scale.

## Methods

### Ethical considerations

The study was approved by the Kenya Medical Research Institute - Scientific and Ethical Review Unit (SSC #3044) and CDC Institutional Review Board (#6806) to use pre-existent, pseudonymized specimens, and data. All individuals, parents and guardians gave written informed consent for themselves or their children to participate in the original studies^23^. For older children aged 13–17 years, assent was obtained as part of the individual informed consenting process.

### Surveillance sites

This study utilised samples collected throughout the surveillance period covering 1^st^ January to 31^st^ December 2014 from three health facility-based respiratory virus surveillance programs in Kenya (Table 1 and Figure 1). The first program included eight inpatient hospitals participating in the influenza sentinel surveillance : Dadaab Refugee Camp (RC), Kakuma RC, Kenyatta National Hospital (KNH), Nakuru County Referral Hospital (CRH), Kakamega CRH, Nyeri CRH, Siaya CRH and Coast General Teaching and Referral Hospital^24–26^; the second included two outpatient clinics; Lwak Mission Hospital, Asembo and Tabitha Medical Clinic, Kibera under Population-Based Infectious Disease Surveillance (PBIDS)^27^, and the third program was a pediatric inpatient sampling at the Kilifi County Hospital (KCH)^28^. The influenza sentinel surveillance was established by Kenya Medical Research Institute-Centre for Global Research (KEMRI-CGHR), the US Centers for Disease Control and Prevention (CDC)-Kenya, and the Ministry of Health as part of the Global Influenza Surveillance and Response System since 2006^24^. The PBIDS platform in Asembo and Kibera is run by KEMRI-CGHR with financial and technical support from CDC since 2006^29^. Surveillance at KCH is conducted by the KEMRI-Centre for Geographic Medical Research Coast (CGMRC) under KEMRI-Wellcome Trust Research Programme (KWTRP) in Kilifi, Kenya^28^. Overall, the selected surveillance sites offer a good representation of the various geographical settings in Kenya.

**Figure 1:**
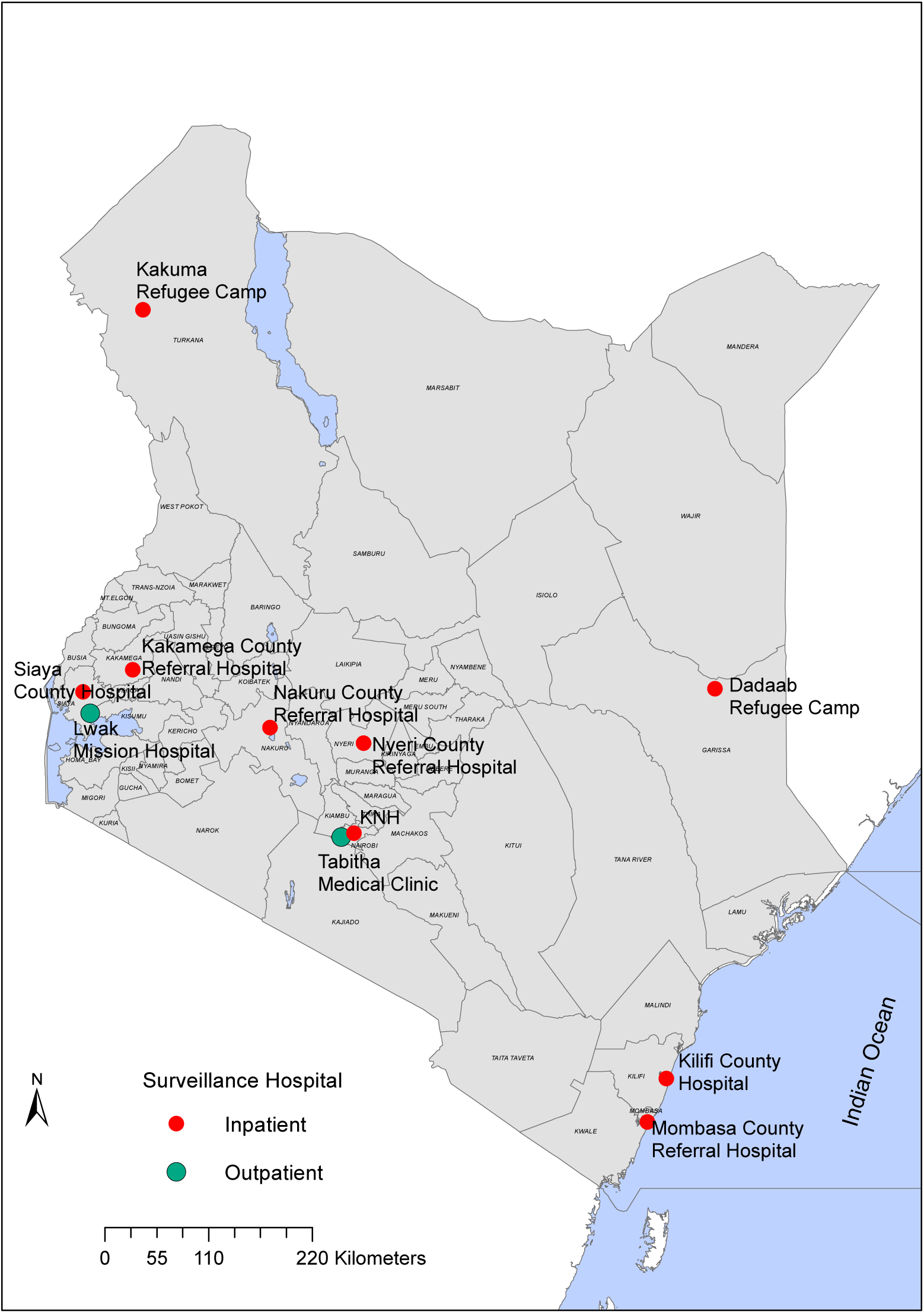
Map of Kenya showing the geo-location of the 11 respiratory surveillance sites, distinguishing inpatient (red markers) from outpatient (green markers) facilities.

**Table 1:**
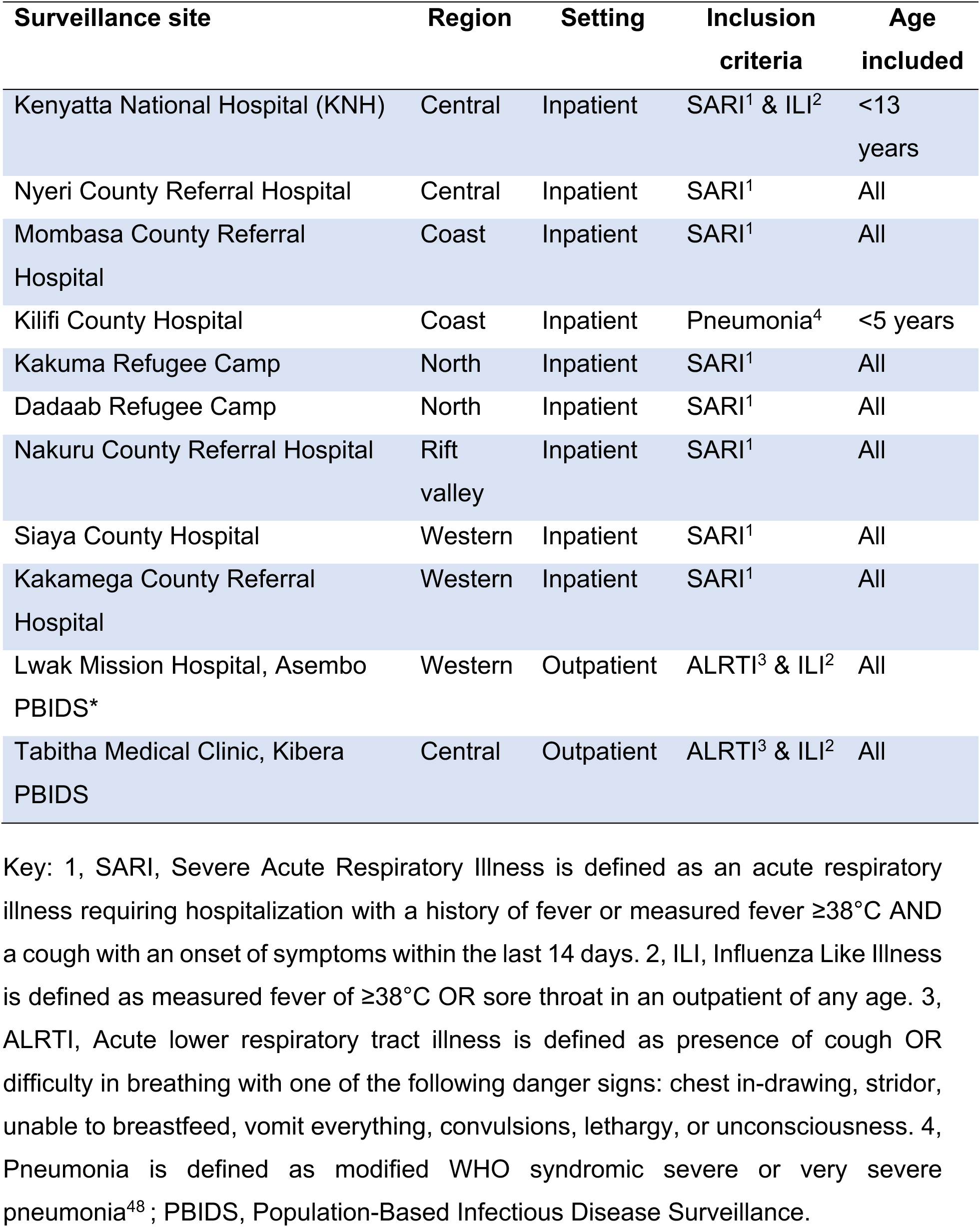
Description of the 11 respiratory surveillance sites across Kenya.

### Patient enrollment

The surveillance sites recruited patients of various ages presenting with acute respiratory illness (ARI) with a measured fever of ≥38°C and a cough with an onset of symptoms within the last 7 days (influenza-like illness, ILI) or acute respiratory illness requiring hospitalization with a history of fever or measured fever ≥38°C and a cough with an onset of symptoms within the last 14 days (severe acute respiratory illness, SARI), or acute lower respiratory tract illness (ALRTI) defined as the presence of cough OR difficulty in breathing with one of the following danger signs: chest in-drawing, stridor, unable to breastfeed, vomit everything, convulsions, lethargy, or unconsciousness or an adaptation of WHO severe/very severe pneumonia as described in a **Table 1**. The different case definitions were consistently applied within each platform over the course of the study period.

### Data and specimen collection

Patient demographic data and clinical features of presenting illness were collected in real-time using custom designed databases. Nasopharyngeal (NP) and/or oropharyngeal (OP) swabs were collected from eligible patients who presented to health facilities with clinical features of an acute respiratory illness as describe in patient enrolment section. NP/OP samples were stored in viral transport media, and transported to the laboratory for long-term storage at −80°C.

### RV screening and sequencing

Viral RNA was extracted using RNeasy Mini Kit (Qiagen, Germany) in Qiacube HT as per the manufacturer’s instructions and screened for respiratory viruses (i.e. RSV (groups A and B), rhinovirus, human coronaviruses (hCoV-OC43, -NL63, -229E), influenza A virus and adenovirus) using a multiplex real-time reverse-transcription PCR (rt-RT-PCR) assay^30, 31^. A sample was considered RV positive if the rt-RT-PCR cycle threshold (Ct) was ≤35.0^32^. The VP4/VP2 viral genomic region (∼420 nucleotides long) of positive samples was amplified and sequenced as previously described^33^. Consensus sequences were assembled using the Sequencher software version 5.4.6 (Gene Codes Corporation, Ann Arbor, USA).

### RV species and type assignment

We used the term ‘type’ to refer to RV sequences classified as distinct by genetic comparisons as described previously^11^. Sequences were assigned into the same RV type based on >90% nucleotide similarity to rhinovirus prototype sequences (also referred to as reference sequences^11^ and phylogenetic clustering with bootstrap support value >70%^11^. Distributions of pairwise genetic distances were assessed for evaluation of intertype and intra-type divergence^11^. Intra-type ‘variant’ was defined based on a divergence cut-off or threshold value determined as the least frequent value between the first (same phylogenetic clade) and second (different phylogenetic clades) modes in a pairwise nucleotide difference distribution plot as previously described^34^. Viruses within the same phylogenetic clade were assumed to belong to the same variant of an RV type.

### Global dataset

Rhinovirus VP4/VP2 sequences from other regions around the world deposited in GenBank as of 31 December 2021, whose sequenced regions overlapped the 803 Kenyan sequences generated in this study and derived from viruses sampled between 1^st^ January 2005 and 31^st^ December 2014, were collated and phylogenetically compared with the Kenya virus sequences. The complete comparison global data set comprised 4,448 VP4/VP2 sequences from 40 countries, including 653 sequences from Kenya (Kilifi (n=627), Nairobi (n=10), Mombasa (n=4), Malindi (n=3), Alupe (n=3), Kisumu (n=2), Isiolo (n=2), Kisii (n=1) and Kericho (n=1) (Appendix Table 1).

### Phylogenetic analysis

Multiple sequence alignments (MSA) were generated using MAFFT v7.220^35^ and maximum likelihood phylogenetic trees estimated using IQ-TREE v1.6.12^36^. Branch support was assessed by 1000 bootstrap iterations. The temporal signal in the data was examined using TempEst v1.5.3^37^. TreeTime v.0.8.1 was used to transform the ML tree topologies into dated trees. Phylogenetic trees were visualized using ggtree v2.2.4 package^38^ in R^39^.

### Statistical analysis

Statistical analysis was conducted using STATA version 15.1 (College Station, Texas). Site-specific monthly percent virus positivity was computed.

### Type diversity calculation

RV type diversity was calculated using diversity indices (Shannon (H) and Simpson (D)) ^40, 41^. The calculations were as follows:

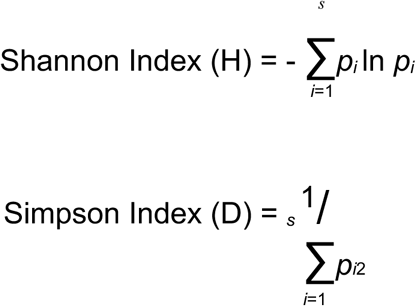

Where *p* is the proportion (n/N) of individual types found in one individual species (n) divided by the total number of types found (N), and s is the total number of species. Higher scores of H and D indicates high diversity.

## Results

Between 1^st^ January and 31^st^ December 2014, a total of 6398 NP and/or OP swabs samples were collected from patients who voluntarily presented to the eleven facilities with ARI. Testing was done on 5859 (91.6%) available samples, Table 2. Of the tested samples, 5665 (96.7%) were linked with their respective demographic and clinical data. Two hundred and fourteen samples (214) collected from Siaya and Kibera patients with missing data on respiratory symptoms were excluded from the study, the final analytical dataset comprised of 5451 specimens. RV was detected in 17.0% (924/5451) of samples collected from the 11 surveillance sites (Table 2). The percent of samples that were virus positive for RV infections varied by surveillance site; Siaya recorded the highest positives (24.0%) while Dadaab (10.4%) recorded the lowest (Table 2). Rhinovirus was detected throughout the year in most sites, although the prevalence of detections varied by month of sampling due to seasonal variation in SARI (Figure 2). Different sites experienced peak occurrence in different months, but a majority saw infection peaks between January - March (7/11) and June - July (5/11) (Figure 2). There was no RV detected in April in Kenyatta National Hospital (KNH) and in May in Kakuma and Kibera sites (Figure 2). There were no samples collected in Dadaab between July and September.

**Figure 2:**
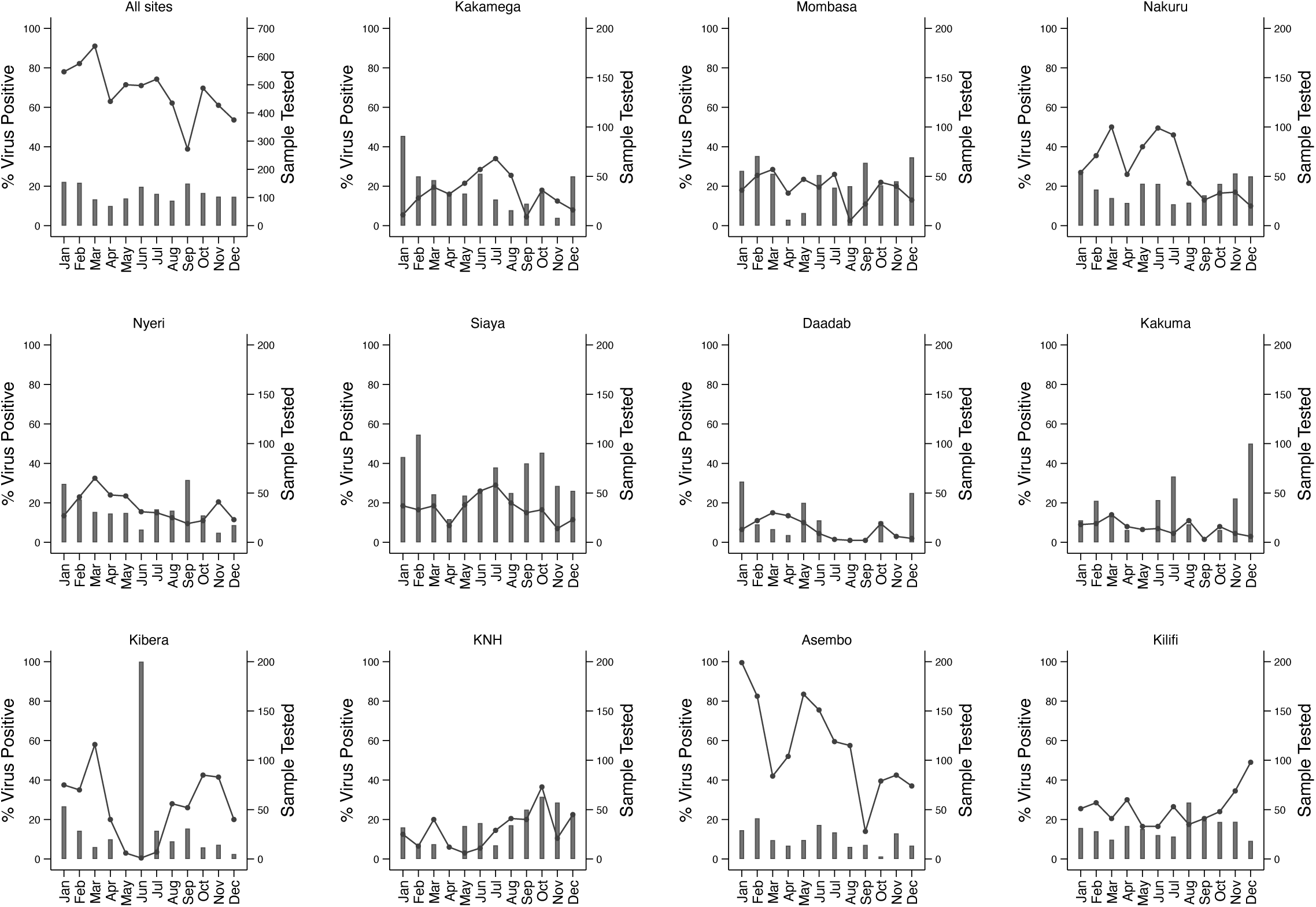
Monthly distribution of samples tested and cases positives for rhinovirus from the 11 surveillance sites in Kenya in 2014.

**Table 2:** Description of samples collected per site, cases positives for RV, and samples sequenced in the VP4/VP2 coding region.

| Surveillance site | NP/OP collections | Tested | Linkage with clinical and demographic data | With respiratory symptoms | Samples positive for HRV | % Positivity | VP4/VP2 sequenced (%) |
| --- | --- | --- | --- | --- | --- | --- | --- |
| Kenyatta National Hospital (KNH) | 510 | 357 | 343 | 343 | 64 | 18.7 | 63 (99) |
| Nyeri County Referral Hospital | 482 | 427 | 425 | 425 | 68 | 16.0 | 57 (84) |
| Mombasa County Referral Hospital | 524 | 455 | 445 | 445 | 103 | 23.1 | 87 (85) |
| Kilifi County Hospital | 722 | 722 | 722 | 722 | 113 | 15.7 | 96 (85) |
| Kakuma Refugee Camp | 220 | 175 | 175 | 175 | 25 | 14.3 | 23 (92) |
| Dadaab Refugee Camp | 189 | 158 | 154 | 154 | 16 | 10.4 | 13 (81) |
| Nakuru County Referral Hospital | 811 | 714 | 712 | 712 | 126 | 17.7 | 103 (82) |
| Siaya County Referral Hospital | 936 | 922 | 794 | 642 | 154 | 24.0 | 128 (83) |
| Kakamega County Referral Hospital | 464 | 418 | 402 | 402 | 72 | 17.9 | 65 (91) |
| Lwak Mission Hospital, Asembo PBIDS* | 879 | 862 | 862 | 862 | 117 | 13.6 | 108(92) |
| Tabitha Medical Clinic, Kibera PBIDS | 640 | 631 | 631 | 569 | 66 | 11.6 | 60 (91) |
| **Not recorded** | 21 | 18 | 0 | 0 | 0 | 0 | 0 |
| **Total** | **6398** | **5859** | **5665** | **5451** | **924** | **17.0** | **803 (86.0)** |

Abbreviations : PBIDS, Population-Based Infectious Disease Surveillance.

VP4/VP2 sequences were successfully obtained from 803/924 (87.0%) RV positive samples (Table 2). The remaining samples either failed to amplify with the VP4/VP2 specific primers (n=111) or were identified as non-RV enteroviruses (n=10). Overall, 492 (61.3%) sequences were classified as *Rhinovirus A* comprising 58 types; 63 (7.8%) sequences were *Rhinovirus B* comprising 16 types, and 248 (30.9%) were *Rhinovirus C* comprising 40 types. The most detected types were A34 (n = 35), A22 (n=30), A58 (n = 27), A12 (n = 25) and A78 (n = 23) (Supplementary Table 1). The number of types largely reflected the number of samples sequenced and varied between sites (Supplementary Figure 1A). Comparison by care setting (i.e., outpatient vs. inpatient) or ARI definition (i.e., ILI vs. SARI vs. ALRTI) across different RV types revealed no significant differences in ARI definition (p-value 0.06) and care setting (p-value 0.2) (Supplementary Table 2). Rhinovirus type diversity per site calculated using the Shannon (H) and Simpson (D) indices indicated that Siaya had the highest value of diversity, followed by Lwak and Kilifi. Dadaab reported the lowest value of diversity (Table 3). All three RV species were detected in all sites, except for Dadaab, where *Rhinovirus B* was not detected (Supplementary Figure 1B). The proportions of RV species were similar in all the sites; *Rhinovirus A* (range, 51 - 75%) and *Rhinovirus C* (range, 17 - 46%) were frequently detected, while *Rhinovirus B* (range, 0 - 13%) infections were low or not detected (Supplementary Figure 1B).

**Table 3:** Rhinovirus type diversity measured using Shannon (H) and Simpson (D) diversity indices.

| Site | Number of sequences | Sequences per species |  |  | Number of types | Shannon's diversity (H) | Simpson diversity (G) |
| --- | --- | --- | --- | --- | --- | --- | --- |
|  |  | RV-A | RV-B | RV-C |  |  |  |
| KNH | 63 | 32 | 2 | 29 | 32 | 3.31 | 23.68 |
| Nyeri | 57 | 31 | 3 | 23 | 35 | 3.38 | 24.43 |
| Mombasa | 87 | 54 | 3 | 30 | 42 | 3.58 | 30.82 |
| Kilifi | 96 | 54 | 7 | 35 | 46 | 3.69 | 34.79 |
| Kakuma | 23 | 14 | 1 | 8 | 15 | 2.56 | 10.76 |
| Dadaab | 13 | 7 | 0 | 6 | 9 | 2.09 | 7.20 |
| Nakuru | 103 | 61 | 7 | 35 | 50 | 3.66 | 28.74 |
| Siaya | 128 | 90 | 16 | 22 | 56 | 3.79 | 34.66 |
| Kakamega | 65 | 49 | 4 | 12 | 33 | 3.29 | 22.59 |
| Asembo | 108 | 65 | 14 | 29 | 52 | 3.74 | 34.23 |
| Kibera | 60 | 33 | 5 | 22 | 38 | 3.47 | 26.17 |

### Spatial-temporal distribution of RV types in Kenya

We identified up to 49 unique RV types co-circulating in a single month across the country, and/or up to 20 unique types within a single location in a single month (Supplementary Figure 2). The duration of circulation varied by RV type; several types occurred at a high prevalence and for consecutive months, while others occurred once or intermittently during the study period (Supplementary Figure 3). For example, RV-A22 was detected throughout the year, A34 was present for 11 consecutive months (February to December 2014), and RV-C7 and A21 circulated consecutively for 8 months (February to September and May to December, respectively) (Supplementary Figure 3). Spatially, several RV types circulated widely; some circulated in multiple sites during the same timeframe, while others circulated in multiple sites at different times (Figure 3). For example, RV-A22 was detected in all 11 sites, while A34 was detected in 10 sites. We observed the occurrence of localized type-specific epidemics or outbreaks; the distribution of RV types was similar between neighbouring locations (Figure 3). For example, types A21 and A78 circulated in Asembo, Siaya, and Kakamega in the same period (June-July), while A49 was seen in Mombasa and Kilifi between July and December (Figure 3). Other types were more random in occurrence with no discernable temporal or spatial pattern.

**Figure 3:**
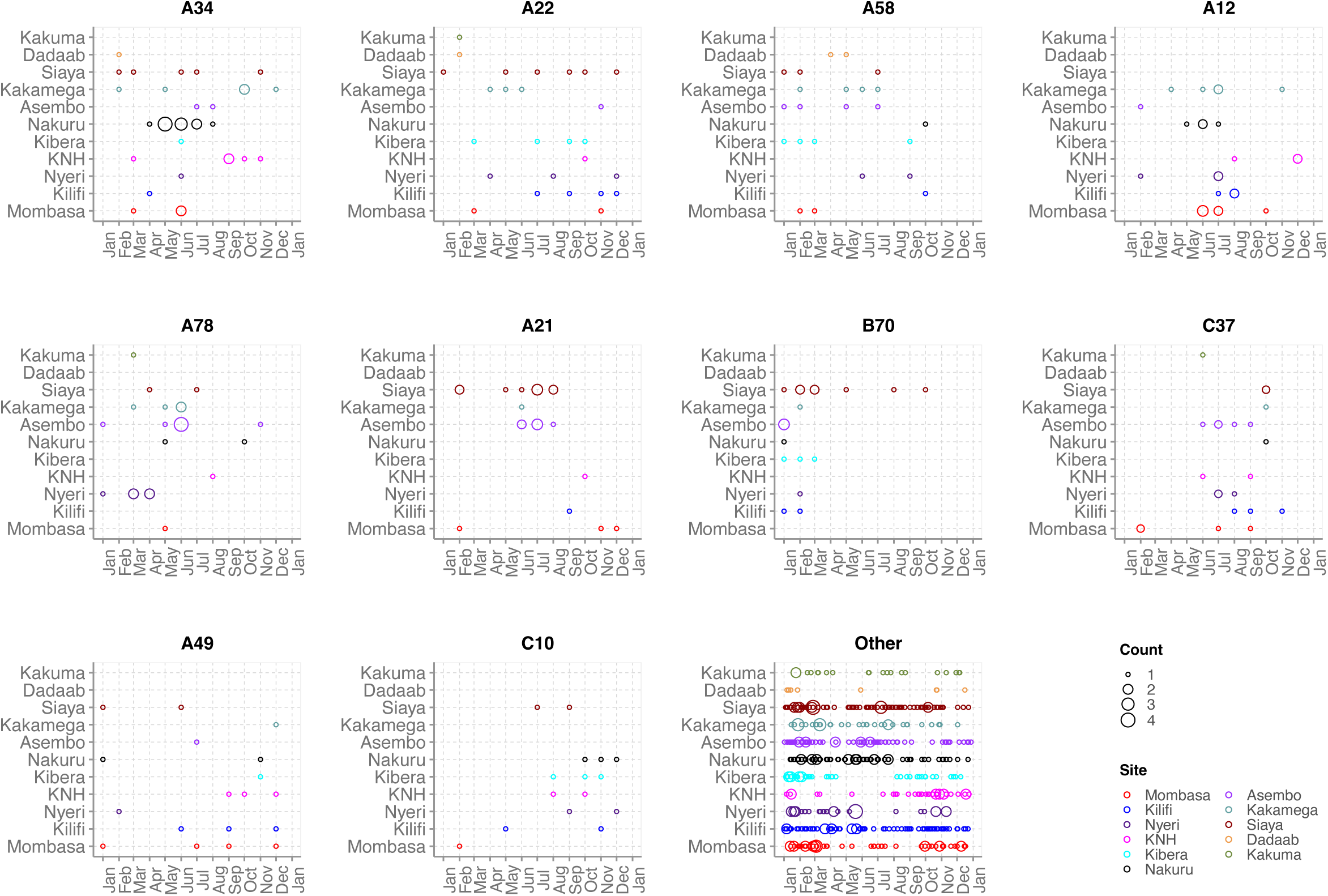
Temporal distributions of frequent RV types across different sites.

### Phylogenetic clustering

We reconstructed time-scaled phylogenies for thirteen prevalent types. In the global context, Kenyan viruses formed monophyletic clusters/clades containing sequences from different sampling sites in Kenya, suggesting multiple introductions and local transmission chains (Figure 4). For example, the Kenyan RV-A22 viruses collected between 2008 and 2014 separated into 3 major clusters, each comprising sequences from different locations in Kenya. Similarly, A34 viruses collected in 2014 separated into 2 major clusters, each with sequences from different sites (Figure 4). Similar observations were observed for other RV types including, A12, A49, A58, A78, and C10 (Figure 4). Certain clades or clusters were location-specific and genetically distinct from other Kenyan 2014 sequences. For example, an A78 variant from Nyeri and an A58 variant in Dadaab (Figure 4), showing evidence of localised transmission clusters.

**Figure 4:**
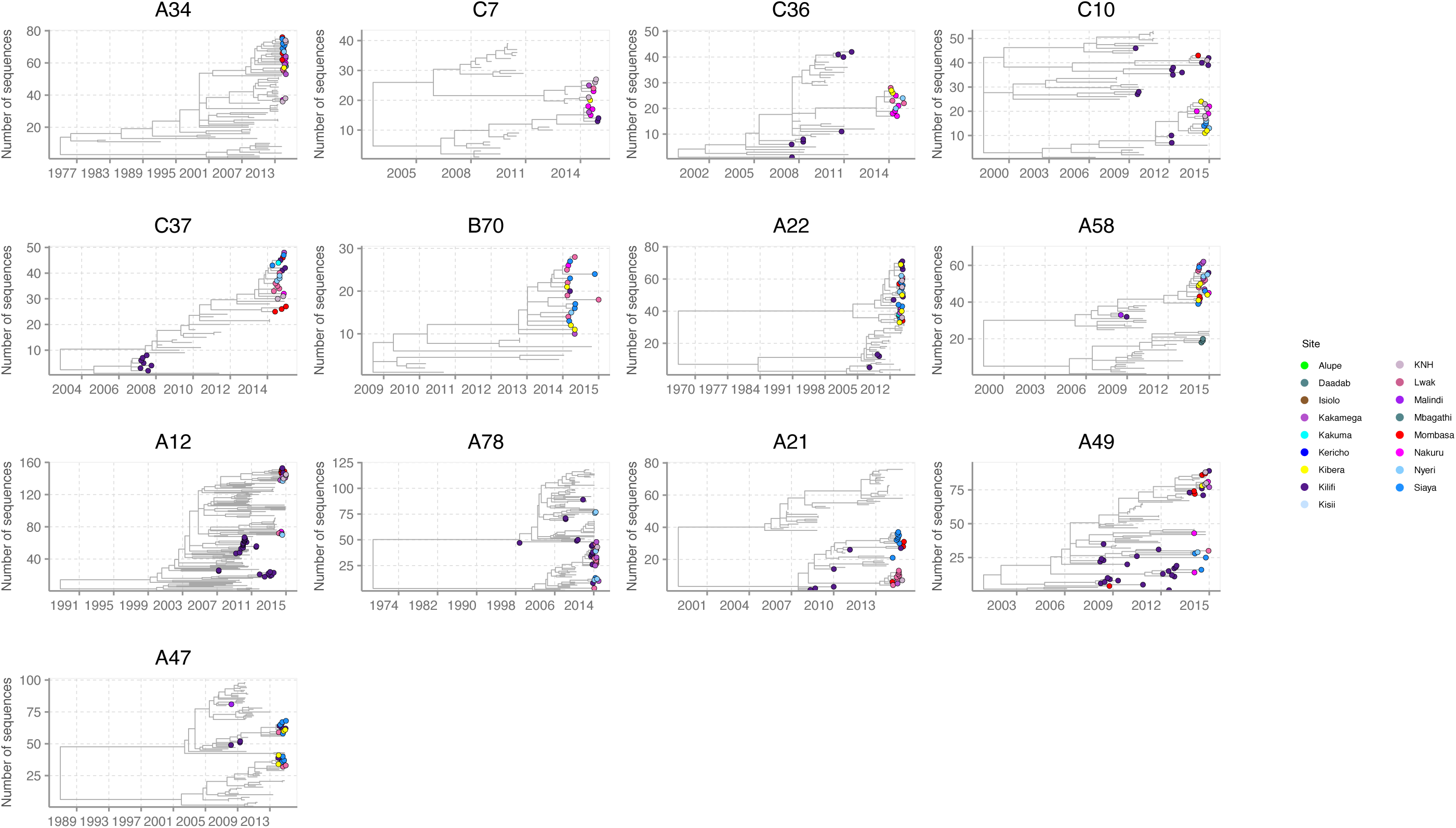
Time-resolved lineage-specific phylogenetic trees for prevalent RV types. The Kenya genomes are indicated with filled circles coloured by site.

Sequences originating from Kenya did not form a single monophyletic group on the phylogenetic tree, instead, they were interspersed mostly as singleton or clusters located separately or with clusters of viruses from other countries. For example, RV-A22 sequences from Kenya clustered closely with sequences from China and France in 2014, while A12 sequences clustered closely with sequences from China, the USA, France, Uganda, and Nepal in 2014. The Kenyan A58 sequences clustered closely with those from USA and France, while the A78 sequences clustered closely with Chinese and American sequences, and the A34 resembled Nepalese sequences.

## Discussion

This study provides a detailed description of the spatial-temporal dynamics of introductions and spread of rhinovirus in Kenya based on phylogenetic analyses. We show that the persistent circulation of RV in Kenya was mostly driven by multiple introductions of different types throughout the one-year period, leading to established local transmission.

*Rhinovirus A* was the predominant species circulating in Kenya in 2014, followed by *Rhinovirus C,* and the least common was *Rhinovirus B*. This is consistent with a previous surveillance study across 8 sampling sites in Kenya in 2008^42^. Although differences in subject recruitment strategies may not allow one-on-one comparison between this study and others, these observations are similar to studies reported in Africa, USA, Asia, Europe, the Middle East, and Australia^11, 43^. The similarities in frequencies of RV species in diverse geographies would suggest that RV circulates unrestricted globally.

RV was detected all year, with the highest detection rates observed in January to March and June to July. The year-round occurrence of rhinovirus in the country was largely sustained by contemporaneous and successive mini epidemics or outbreaks caused by distinct types and variants that were probably introduced separately into the country or diversified locally after a single introduction.

Spatial-temporal analysis revealed occurrence of multiple types and variants across space and time in agreement with previous studies^12, 14, 44^. The observed variations in time and duration of circulation among the rhinovirus types could be due to (i) differences in the duration of type-specific immunity, (ii) frequency of introductions or, iii) level of antigenic similarity (heterologous immunity)^34^. The concurrent and sequential circulation of RV types seen as the occurrence of multiple peaks of the same type in the same or distant locations could signify multiple introductions, antigenic variation, or infections in different population strata^34^. The heterogeneity in RV detection by month and location could be attributed to the seasonal variation in SARI or sampling methods or to regional differences in environmental and climatic factors. Climatic factors such as temperature, rainfall, and relative humidity have been hypothesized to influence RV activity in the tropics^3, 45, 46^.

Even with non-uniform sampling and short duration of study (1 year), we showed close genetic association among sequences from different sites as a result of widespread transmission of the virus in the country. On a finer spatial scale, study sites occurring within the same geographical region (as described in Table 1) had a similar distribution of RV types, an indicator of a point source outbreak, in which a single virus type enters a location and diffuses through the interconnected populations probably sharing social amenities^12^. This agrees with our previous work that showed greater similarity in RV types among the locations in proximity^12^.

The global phylogenies showed that RV viruses circulating in Kenya were closely related to strains circulating in Europe, Asia, and North America. The Kenyan RV diversity appears to be nested within the global diversity as a result of transmission facilitated by unrestricted movement, increased connectivity, and social mixing.

Although our analyses were limited to the VP4/VP2 genomic region, we highlight the use of sequence data to trace the introduction and spread of rhinovirus at the countrywide level and show the benefit of systematic, continuous, and geographically representative surveillance to detect and monitor the occurrence of types at a larger scale. Whole genome sequencing could provide more insight into virus diversity and transmission^47^, and we recommend that future studies should combine genomic data with epidemiological and anthropological data (e.g., host migration, immunity profiles, population densities, and social contact patterns) to further elucidate patterns of RV infections.

This study had some limitations. First, due to the retrospective approach of the study, it was not possible to recover sequences from all the samples due to sample degradation; samples (n=111/924) that failed sequencing had considerably higher Ct values (low viral load) compared to samples that were successfully sequenced with low Ct values (high viral load). Second, there were varying study design by sites, for instance, the disease case definition and ages enrolled. The varying designs would mainly affect the virus positivity estimates observed over the one-year surveillance period, in that sites that had more inclusive case definition that includes mild cases of respiratory illnesses may exhibit higher rates of rhinovirus positivity compared to sites using a more restrictive case definition that only includes severe cases of respiratory illnesses. Third, the inclusion of fever as a criterion in most of the case definitions used may lead to the underestimation of RV prevalence since many RV infections, including severe cases, do not necessarily manifest with fever. Consequently, there is a possibility that we might have missed RV patients who do not exhibit fever symptoms, potentially impacting the overall assessment of RV prevalence in our study. Fourth, the short sequence fragment analysed in this study may result in spurious phylogenetic connections. Lastly, the study focused on samples that were collected in hospital settings, which may have missed genotypes associated with asymptomatic or mildly ill infections.

In conclusion, our study demonstrates that sustained circulation of the virus in 2014 was due to frequent introductions of different types and variants into the country followed by local spread for some of these introductions. Temporal differences in persistence of rhinovirus types over the one-year period, could be attributed to differences in the frequency and number of virus introductions into the country. Spatial patterns show extensive spread of the virus, and the evidence of similar distribution of types in locations that are in proximity to each other may indicate local partitioning or spatial structures of virus transmission.

## Supporting information

manuscript

## Acknowledgements

We thank all the study participants for their contribution of study specimens and data and surveillance staff from all the study sites included. We acknowledge the laboratory staff at KEMRI-Wellcome Trust Research Programme and CDC-supported laboratory at KEMRI-Centre for Global Health Research for specimen handling and testing. The primary data and specimen collection in Influenza surveillance and PBIDS sites were supported by U.S. Centers for Disease Control and Prevention while pediatric surveillance at Kilifi County Hospital was funded by the Wellcome Trust.

## Author Contributions

J.M.M, E.K., C.A., PKM, D.J.N., Conceptualised and designed the study

J.M.M.,M.M.L, C.L., M.M., Provided laboratory sequencing and support

J.M.M., M.M.L, E.K., C.A., D.J.N., Provided phylogenetic analysis

J.M.M., N.M. Provided data curation and analysis

J.M.M., E.K., C.A., D.J.N., Wrote the original manuscript draft

All authors read and approved the final manuscript.

## Data availability

Sequence data generated in this study are available in GenBank under accession numbers: MZ129390 - MZ130096. Additional data and analysis scripts for this manuscript are available at the VEC Harvard Dataverse: https://doi.org/10.7910/DVN/CBHVTA

## Disclaimer

The findings and conclusions in this report are those of the author(s) and do not necessarily represent the official position of U.S. the Centers for Disease Control and Prevention.

