## Supplementary material for "Spatio-temporal distribution of rhinovirus types in Kenya: A retrospective analysis, 2014": manuscript

**Supplementary Table 1:** Rhinovirus types identified in Kenya in 2014

| | | | | | | | | | | | | | | | | | | | |
|---|---|---|---|---|---|---|---|---|---|---|---|---|---|---|---|---|---|---|---|
| **RV-A** | | | | | | | | | | | | | | | | | | | |
| **A34** | **A22** | **A58** | **A12** | **A78** | **A21** | **A49** | **A61** | **A81** | **A54** | **A20** | **A82** | **A10** | **A47** | **A55** | **A63** | **A103** | **A106** | **A13** | **A31** |
| 35 | 30 | 27 | 25 | 23 | 21 | 20 | 16 | 16 | 15 | 14 | 12 | 11 | 11 | 10 | 10 | 9 | 9 | 9 | 9 |
| **A43** | **A89** | **A38** | **A90** | **A29** | **A33** | **A94** | **A53** | **A80** | **A104** | **A16** | **A28** | **A65** | **A68** | **A11** | **A18** | **A23** | **A40** | **A1** | **A101** |
| 9 | 9 | 8 | 8 | 7 | 7 | 7 | 6 | 6 | 5 | 5 | 5 | 5 | 5 | 4 | 4 | 4 | 4 | 3 | 3 |
| **A30** | **A36** | **A39** | **A46** | **A51** | **A105** | **A59** | **A62** | **A96** | **A102** | **A15** | **A2** | **A45** | **A56** | **A67** | **A77** | **A8** | **A88** | | |
| 3 | 3 | 3 | 3 | 3 | 2 | 2 | 2 | 2 | 1 | 1 | 1 | 1 | 1 | 1 | 1 | 1 | 1 | | |
| **RV-B** | | | | | | | | | | | | | | | | | | | |
| **B70** | **B92** | **B26** | **B4** | **B42** | **B27** | **B35** | **B91** | **B102** | **B17** | **B3** | **B69** | **B79** | **B83** | **B93** | **B97** | | | | |
| 19 | 9 | 8 | 5 | 5 | 3 | 3 | 2 | 1 | 1 | 1 | 1 | 1 | 1 | 1 | 1 | | | | |
| **RV-C** | | | | | | | | | | | | | | | | | | | |
| **C37** | **C10** | **C46** | **C27** | **C14** | **C7** | **C36** | **C15** | **C43** | **C18** | **C35** | **C40** | **C47** | **C16** | **C25** | **C42** | **C48** | **Cpat19** | **C12** | **C1** |
| 22 | 18 | 15 | 14 | 13 | 13 | 12 | 10 | 9 | 8 | 8 | 8 | 8 | 6 | 6 | 6 | 6 | 5 | 5 | 4 |
| **C26** | **C3** | **C33** | **C6** | **C9** | **C23** | **C31** | **C39** | **C17** | **C21** | **C28** | **C29** | **C49** | **Cpat27** | **C11** | **C13** | **C19** | **C22** | **C30** | **C38** |
| 4 | 4 | 4 | 4 | 4 | 3 | 3 | 3 | 2 | 2 | 2 | 2 | 2 | 1 | 1 | 1 | 1 | 1 | 1 | 1 |

**Supplementary Table 2:** Comparative analysis of ARI definitions and care settings across the three rhinovirus species (A, B, and C).

| | RV Species | | | | |
|---|---|---|---|---|---|
| | **RV A** | **RV B** | **RV C** | **Total** | **p-value** |
| **ARI definition** | | | | | |
| **ALRTI & ILI** | 87 (17.68%) | 17 (26.98 %) | 44 (17.74%) | 148 | 0.06 |
| **SARI & ILI** | 33 (6.71%) | 1 (1.59%) | 28 (11.29%) | 62 | |
| **SARI** | 317 (64.43) | 37 (58.73 %) | 140 (56.45%) | 494 | |
| **Severe/VSevere Pneumonia** | 55 (11.18%) | 8 (12.69%) | 36 (14.52%) | 99 | |
| **Care setting** | | | | | |
| **Outpatient** | 87 (17.68%) | 17 (26.98%) | 44 (17.74%) | 148 | 0.27 |
| **Inpatient** | 405 (82.31%) | 46 (73.01% | 204 (82.26%) | 655 | |

Abbreviation: ARI, Acute respiratory illness

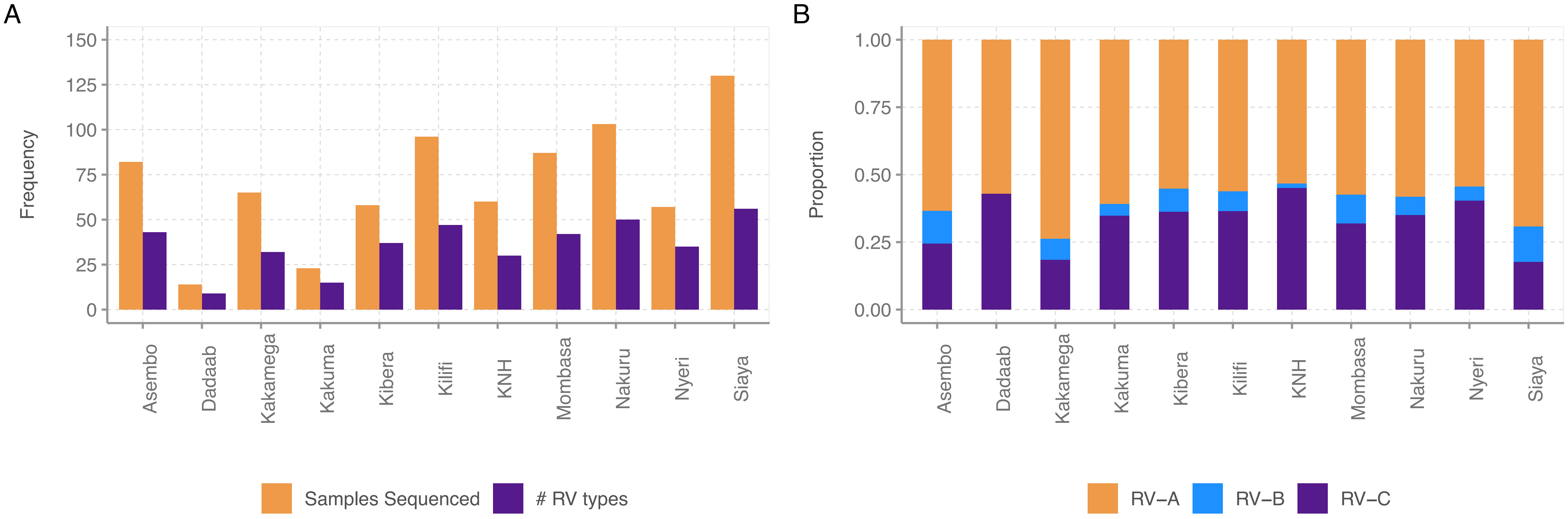

**Supplementary Figure 1:** (A) Number of samples sequenced per site versus number of types recorded B) Proportion of rhinovirus (RV) species detected per site

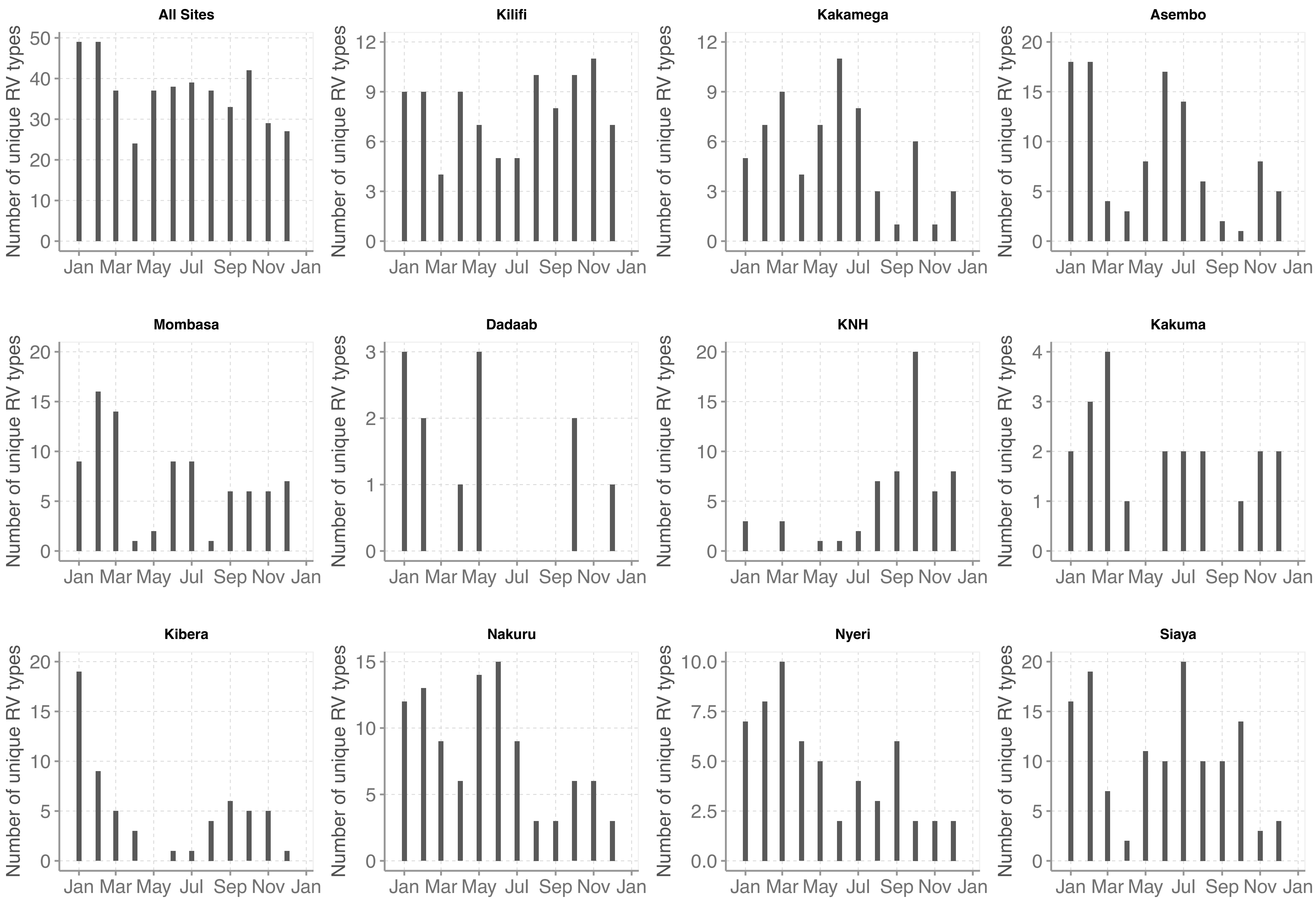

**Supplementary Figure 2:** Monthly frequency of RV types detected per study site

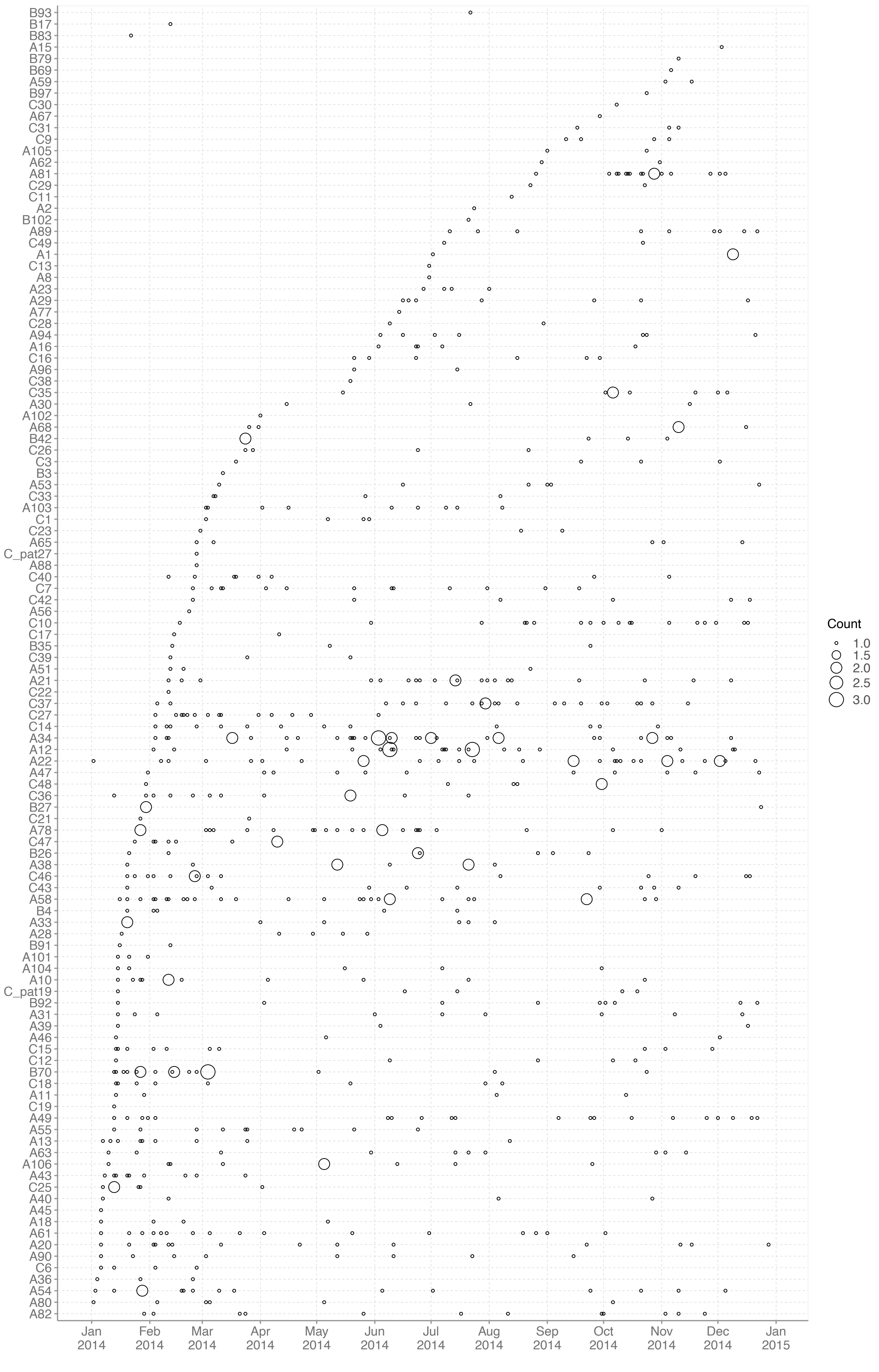

**Supplementary Figure 3:** Monthly temporal occurrence plots for prevalent RV types
